# Mapping inherited genetic variation with opposite effects on autoimmune disease and cancer identifies candidate drug targets associated with the anti-tumor immune response

**DOI:** 10.1101/2023.12.23.23300491

**Authors:** Junyu Chen, Michael P. Epstein, Joellen M. Schildkraut, Siddhartha P. Kar

## Abstract

**Background:** Germline alleles near genes that encode certain immune checkpoints (*CTLA4*, *CD200*) are associated with autoimmune/autoinflammatory disease and cancer but in opposite directions. This motivates a systematic search for additional germline alleles which demonstrate this pattern with the aim of identifying potential cancer immunotherapeutic targets using human genetic evidence.

**Methods:** Pairwise fixed effect cross-disorder meta-analyses combining genome-wide association studies (GWAS) for breast, prostate, ovarian and endometrial cancers (240,540 cases/317,000 controls) and seven autoimmune/autoinflammatory diseases (112,631 cases/895,386 controls) coupled with *in silico* follow-up. To ensure detection of alleles with opposite effects on cancer and autoimmune/autoinflammatory disease, the signs on the beta coefficients in the autoimmune/autoinflammatory GWAS were reversed prior to meta-analyses.

**Results:** Meta-analyses followed by linkage disequilibrium clumping identified 312 unique, independent lead variants with P_meta_<5×10^-8^ associated with at least one of the cancer types at P_cancer_<10^-3^ and one of the autoimmune/autoinflammatory diseases at P_auto_<10^-3^. At each lead variant, the allele that conferred autoimmune/autoinflammatory disease risk was protective for cancer. Mapping each lead variant to its nearest gene as its putative functional target and focusing on genes with established immunological effects implicated 32 of the nearest genes. Tumor bulk RNA-Seq data highlighted that the tumor expression of 5/32 genes (*IRF1*, *IKZF1*, *SPI1*, *SH2B3*, *LAT*) were each strongly correlated (Spearman’s ρ>0.5) with at least one intra-tumor T/myeloid cell infiltration marker (*CD4*, *CD8A*, *CD11B*, *CD45*) in every one of the cancer types. Tumor single-cell RNA-Seq data from all cancer types showed that the five genes were more likely to be expressed in intra-tumor immune versus malignant cells. The five lead SNPs corresponding to these genes were linked to them via expression quantitative trait locus mechanisms and at least one additional line of functional evidence. Proteins encoded by the genes were predicted to be druggable.

**Conclusion:** We provide population-scale germline genetic and functional genomic evidence to support further evaluation of the proteins encoded by *IRF1*, *IKZF1*, *SPI1*, *SH2B3*, and *LAT* as possible targets for cancer immunotherapy.

## Introduction

Immunotherapy of cancer through inhibition of the immune checkpoints CTLA-4 and PD-1/PD-L1 has led to dramatic improvements in survival for patients who respond to these treatments across several cancer types. An emerging body of evidence suggests that inherited or germline genetic variation with established association with autoimmune disease susceptibility, when aggregated into multi-variant polygenic scores predictive of autoimmune disease, is associated with the risk of developing immune-related adverse events in cancer patients receiving immune checkpoint inhibitors and, in turn, with better therapeutic response and survival likely due to the enhanced autoimmune anti-tumor activity (1,2). Germline single nucleotide polymorphisms (SNPs) known to be associated with autoimmune disease risk have also been shown to associate with intra-tumor immune cell infiltrate levels underscoring the importance of the germline genome in regulating the anti-tumor immune response (3,4).

A SNP rs231779, associated at genome-wide significance (P<5×10^-8^) with rheumatoid arthritis, hypothyroidism, and type 1 diabetes risk, was recently found to be associated with predisposition to melanoma and keratinocyte cancers (5). There are three notable features of this finding: the gene nearest to rs231779 is *CTLA4*, the allele of rs231779 that increases autoimmune disease risk is protective for melanoma and keratinocyte cancers, and rs231779, while being associated with melanoma risk (P=3.6×10^-4^), did not reach the conventional genome-wide significance threshold (P<5×10^-8^). Even more recently, germline alleles near genes encoding the CD200 immune checkpoint – specifically, *CD200*, *CD200R1* that encodes the receptor for CD200, and *DOK2* that encodes a downstream adapter protein – were found to have significant associations with, but opposite effects on, autoimmune hypothyroidism, asthma and eczema versus basal cell carcinoma risks, recapitulating the pattern of pleiotropy observed at the *CTLA4* genomic locus (6). Moreover, tumor *CD200R1* expression was strongly correlated (Spearman’s ρ>0.5) with the expression of at least one of four commonly used T cell and myeloid infiltration markers (*CD4*, *CD8A*, *CD11B*, and *CD45*) in tumors from multiple cancer types in The Cancer Genome Atlas (TCGA; (6)). These observations have formed the basis for the development of an anti-CD200R1 antibody as a CD200 checkpoint inhibitor with a promising pre-clinical profile that is now in Phase 1/2a trials (6).

A vast amount of genome-wide association study (GWAS) data for both autoimmune/autoinflammatory conditions and cancers now exist in the public domain. However, despite the striking pleiotropic pattern of opposite but significant allelic effects exhibited by SNPs near well-established (CTLA-4) and novel (CD200R1) immune-oncology targets, there is no published catalog of SNPs that demonstrate this pattern from systematic mining of these GWAS data. To address this gap and identify potential additional candidates for follow up in immunotherapeutic drug development programs, here we present the results from large-scale pairwise cross-disorder meta-analyses combining GWAS data on 112,631 autoimmune/autoinflammatory disease and 240,540 cancer cases. Our analyses, guided by data availability, focused on seven adult autoimmune/autoinflammatory diseases – type 1 diabetes (T1D), rheumatoid arthritis (RA), Hashimoto’s thyroiditis (HT), multiple sclerosis (MS), systemic lupus erythematosus (SLE), ulcerative colitis (UC) and Crohn’s disease (CD) – and four cancers – breast, prostate, ovarian, and endometrial cancers. We sought to identify independent (in terms of linkage disequilibrium), genome-wide significant lead SNPs in each pairwise meta-analysis where the same allele was nominally associated with an autoimmune/autoinflammatory disease and a cancer type but with the opposite direction of effect and adapted our meta-analytic approach to power the discovery of such SNPs. We mapped each identified lead SNP to its nearest gene as its most likely regulatory target and focused on the nearest genes with known immune system function. We then found five genes among the 32 immune-related nearest genes whose tumor expression was strongly correlated with the tumor expression of at least one T cell or myeloid infiltration marker in all four cancer types (2,696 breast, prostate, ovarian and endometrial tumors). Each of the five genes was, in general, more highly expressed in tumor-infiltrating immune cells versus malignant cells across 29 single-cell RNA sequencing data sets obtained from these four cancer types. We confirmed that the lead SNPs corresponding to the five genes – *IRF1*, *IKZF1*, *SPI1*, *SH2B3* and *LAT* – were expression quantitative trait loci for these genes, allowing us to establish the directionality of the relationship between their expression and cancer risk. We also identified additional functional genomic evidence to consolidate the link between these lead SNPs and the five corresponding genes and determined that the proteins encoded by the genes were potentially “druggable” via antibodies or proteolysis targeting chimeras (PROTACs).

## Methods

### GWAS data sets

We used publicly available GWAS summary statistics for four cancer types and seven autoimmune/autoinflammatory diseases in our study of pleiotropic SNPs across these cancers and autoimmune diseases (Supplementary Table 1). Summary statistics consisted of SNP rs number/identifier, chromosome, position, effect allele, other allele, effect allele frequency, imputation quality score, effect size estimate or beta coefficient from regression models for genetic association, standard error for this estimate and *P*-value. All summary statistics were based on GWAS conducted in individuals of European or predominantly European ancestry and the human genome build for these data was GRCh37. SNPs with minor allele frequency<0.5% and imputation quality score (*r*^2^)<0.3 were excluded, except for the GWAS of CD and UC, which did not report allele frequency and imputation quality metrics in the publicly available data sets.

Breast cancer risk GWAS summary statistics were obtained from meta-analyses of 122,977 overall, 69,501 estrogen receptor (ER)-positive, and 21,468 ER-negative breast cancer cases, with each set of cases compared against 105,974 controls (7). GWAS summary statistics for ovarian cancer risk were obtained from a meta-analysis involving 25,509 cases (that included 13,037 high-grade serous cases) and 40,941 controls (8). GWAS summary statistics were obtained from a meta-analysis of 79,148 cases and 61,106 controls for prostate cancer risk (9); and from a meta-analysis involving 12,906 cases and 108,979 controls for endometrial cancer risk (10).

CD GWAS summary statistics were sourced from a meta-analysis of 4,474 cases and 9,500 controls (11). The same study also reported UC GWAS summary statistics from a meta-analysis involving 4,173 cases and 9,500 controls (11). Summary statistics were obtained from GWAS meta-analyses of 29,880 cases and 73,758 controls for RA (12), 7,219 cases and 15,991 controls for SLE (13), 30,234 cases and 725,172 controls for HT (14), 14,498 cases and 24,091 controls for MS (15), and 22,153 cases and 37,374 controls for T1D (16).

### Meta-analysis for each autoimmune/autoinflammatory disease and cancer pair

We first performed a meta-analysis of each autoimmune/autoinflammatory disease-cancer type or subtype pair using the fixed effect inverse-variance weighted method implemented in METAL (17). Apart from analyzing overall breast, prostate, ovarian and endometrial cancer GWAS we also separately evaluated ER-positive and -negative breast and high-grade serous ovarian cancers since these subtypes had substantial GWAS sample sizes in their own right and are known to have distinct genetic architectures. Thus, we effectively crossed seven autoimmune/autoinflammatory diseases GWAS with seven cancer type/subtype GWAS in the paired meta-analyses. We only included SNPs with P<10^-3^ for association with both cancer *and* autoimmune disease in each pairwise meta-analysis, with the choice of this P<10^-3^ threshold guided by our previously published cross-cancer GWAS meta-analysis (18). In order to obtain meta-analysis results such that SNPs that have opposite effects on the two outcomes (autoimmune/autoinflammatory disease versus cancer type/subtype) would be identified at combined genome-wide significance (P<5×10^-8^) under the fixed effect model, beta coefficients (effect size estimates) from GWAS summary statistics for all the autoimmune diseases were multiplied by -1 before the meta-analysis thus reversing the sign on the estimate while keeping the effect allele the same. In addition, after reversing the sign and performing each pairwise meta-analysis, Cochrane’s Q test for heterogeneity was applied and SNPs with P<0.05 for heterogeneity were excluded from further analysis.

Meta-analysis results from METAL were used as input for the Functional Mapping and Annotation (FUMA) platform to identify lead SNPs associated with both a cancer and an autoimmune/autoinflammatory disease (19) using linkage disequilibrium (LD) clumping. For LD clumping in FUMA, the maximum (largest numeric value) *P*-value of lead SNPs was set as 5×10^-8^; the maximum distance between LD blocks to merge into a single region was set to 1 Mb; and the LD *r*^2^ threshold to define lead SNPs was set to 0.1. The combination of the sign reversal process described above, the application of the heterogeneity test, and the P<10^-3^ individual trait and P<5×10^-8^ cross-trait meta-analysis thresholds collectively ensured that our final set of lead SNPs were (1) nominally associated with at least one autoimmune/autoinflammatory disease and one cancer type/subtype, (2) the direction of allelic effect was opposite between the autoimmune/autoinflammatory disease and cancer, but (3) the allelic effect size was otherwise homogeneous across the traits, and (4) the combined cross-trait association was genome-wide significant.

### Identification of immune-related genes among genes nearest to lead SNPs

Large-scale systematic evaluation of the GWAS literature has suggested that in about 70% of the instances the gene nearest to a GWAS-identified lead SNP was the most likely target gene of that lead SNP locus (20). Therefore, we used the FUMA pipeline to annotate the lead SNPs identified in each pairwise cancer-autoimmune/autoinflammatory disease meta-analysis with its nearest gene. We hypothesized that some, but not necessarily all, of these genes would have functions related to the immune system and to identify this subset of immune-related genes we merged our nearest gene list with a list of 1,793 immune-related genes curated by the ImmPort project (21,22) to select the genes that were in common to both lists. Second, as a strategy to identify additional immune-related genes potentially missed by the ImmPort approach we performed pathway enrichment analysis for the combined list of all nearest genes identified across all the pairwise meta-analyses. We used the Enrichr platform (23) for the pathway analysis, applying the hypergeometric test to identify pathway overrepresentation and four pathway databases to obtain pathway annotations (Reactome 2022, WikiPathway 2021, KEGG 2021 and MSigDB Hallmark 2020). We then assessed the top 10 results from each pathway database, selected the immune-related pathways among the top 10, and picked the nearest genes from our list that were driving that immune-related pathway association as per Enrichr.

### Tumor bulk and single-cell RNA-Seq analyses to prioritize immune-related nearest genes based on association with intra-tumor immune cell infiltration

We downloaded tumor bulk RNA-Seq gene expression data for 1,084 breast, 494 prostate, 589 ovarian and 529 endometrial tumors profiled in The Cancer Genome Atlas (TCGA) PanCancer Atlas project from the cBioPortal (24). Specifically, we used the log-transformed, RNA-Seq by expectation maximization (RSEM V2) and Z-score normalized data with the Z-scores computed relative to all samples. We subset these data to retain the nearest genes prioritized by the ImmPort and/or Enrichr analyses and used the Spearman’s rank correlation coefficient (ρ) to calculate the correlation between the expression of each of these genes and four T lymphocyte or myeloid white blood cell markers (*CD4*, *CD8A*, *CD11B* and *CD45*), the choice of these tumor immune cell infiltrate markers guided by and identical to the markers used by Fenaux, et al. (6). Genes with Spearman’s ρ>0.5 in the correlation analyses for at least one immune cell infiltrate marker across all four cancer types were defined as prioritized genes. Relative cell-specificity in expression of the prioritized genes was evaluated using 12 breast, 6 prostate, 9 ovarian and 2 endometrial tumor single-cell RNA-Seq data sets available in the Tumor Immune Single-cell Hub 2 (TISCH2) database (25). Specifically, we sought evidence that across the four cancer types the prioritized genes had higher expression in tumor infiltrating immune cells as compared to malignant, stromal, or other cell types within the tumor. Collectively, the purpose of these bulk and single-cell RNA-Seq analyses was to establish correlations in expression between the prioritized nearest genes and the intra-tumor immune response.

### Functional annotation to link prioritized genes and their corresponding lead SNPs

For the prioritized genes, we performed lookups of the corresponding lead SNPs in the OpenTargets Genetics database (26). OpenTargets provided information on expression/splice/protein quantitative trait loci (e/s/p-QTLs), promoter capture Hi-C chromosome conformation and chromatin interactions, and variant coding effect predictions (26). This yielded functional genomic evidence that reinforced the status of the nearest gene as the most likely target of the lead SNP for the nearest genes prioritized by the combination of the immune-related gene annotation and intra-tumor bulk RNA-Seq expression immune marker correlation analyses. We also assessed the directionality of our findings using blood-based local or *cis*-eQTL data for 31,684 individuals from the eQTLGen consortium (*cis* being defined as genomic distance<1 megabase between the eQTL SNP and the gene whose expression it regulates; (27)). Specifically, we assessed the direction of regulation of expression for each prioritized gene for the effect allele of the corresponding lead SNP that had opposite effects on autoimmune/autoinflammatory disease and cancer. For genes not available in eQTLGen, we performed a lookup of the Genotype Tissue Expression (GTEx) project whole blood eQTL data (28).

### Evaluating the druggability of proteins encoded by the prioritized genes

We examined the druggability of the proteins encoded by the prioritized genes using the DrugnomeAI tool (29). DrugnomeAI is a stochastic, semi-supervised machine learning framework recently developed to predict the druggability of proteins encoded by the human exome. Within this framework, genes encoding proteins that are established targets of known drugs are labeled and 324 gene-level predictors curated from 15 data sources are modelled using an ensemble of classifiers to generate predictions of whether a particular unlabeled gene/protein is likely to be therapeutically targetable. The DruggnomeAI predictions take the form of percentile scores (higher being better, with scores>90^th^ percentile generally deemed “high probability” candidates) ranking each gene/protein relative to all other genes/proteins for targeting by each of three specific drug modalities: small molecules, antibodies and PROTACs. The framework also provides oncology-specific prediction models for gene/protein targeting by small molecule- and antibody-based drugs.

## Results

Our pairwise cross-disorder meta-analyses followed by linkage disequilibrium-based clumping to identify independent lead SNPs yielded 80 overall breast cancer, 83 ER-positive breast cancer, 35 ER-negative breast cancer, 27 ovarian cancer, 20 high-grade serous ovarian cancer, 101 prostate cancer and 52 endometrial cancer susceptibility alleles that were associated with the cancer at P<10^-3^ and had an opposite effect on, and P<10^-3^ association with, at least one of the seven autoimmune/autoinflammatory diseases (Supplementary Table 2 for total lead SNPs by analysis, Supplementary Tables 3 to 9 for details of lead SNPs from each pairwise meta-analysis). Each of these 398 lead SNPs reached genome-wide significance (P<5×10^-8^) in the pairwise cross-disorder meta-analysis where it was identified and demonstrated little statistical evidence of heterogeneity in effect sizes themselves (Cochran’s Q test P>0.05) though the direction of effect across cancer and autoimmune disease were opposite. These 398 lead SNPs consisted of 312 unique SNPs including 244 that were identified in only one paired cancer-autoimmune/autoinflammatory disease meta-analysis and 68 SNPs that were identified in multiple paired meta-analyses.

Annotating the lead SNPs identified from the pairwise cancer-autoimmune/autoinflammatory disease meta-analyses with their nearest genes using FUMA and then overlapping the nearest gene list with immune-related genes from ImmPort (Supplementary Table 10) and Enrichr (Supplementary Table 11) revealed a total of 32 genes that were a nearest gene *and* a immune-function-related gene (Table 1; 16 ImmPort genes, 20 Enrichr genes; 4 overlapping between the methods). The expression of five of these 32 genes (*IRF1*, *IKZF1*, *SPI1*, *SH2B3* and *LAT*) were strongly correlated (Spearman’s ρ>0.5) with the expression of at least one tumor immune cell infiltrate marker (*CD4*, *CD8A*, *CD11B* and *CD45*) in all four cancer types (breast, prostate, ovarian and endometrial) in bulk RNA-Seq data from TCGA (Figure 1). In fact, all five of these genes had a minimum ρ>=0.37 for all four intra-tumor T lymphocyte/myeloid cell markers across all four cancers (Figure 1).

**Figure 1.**
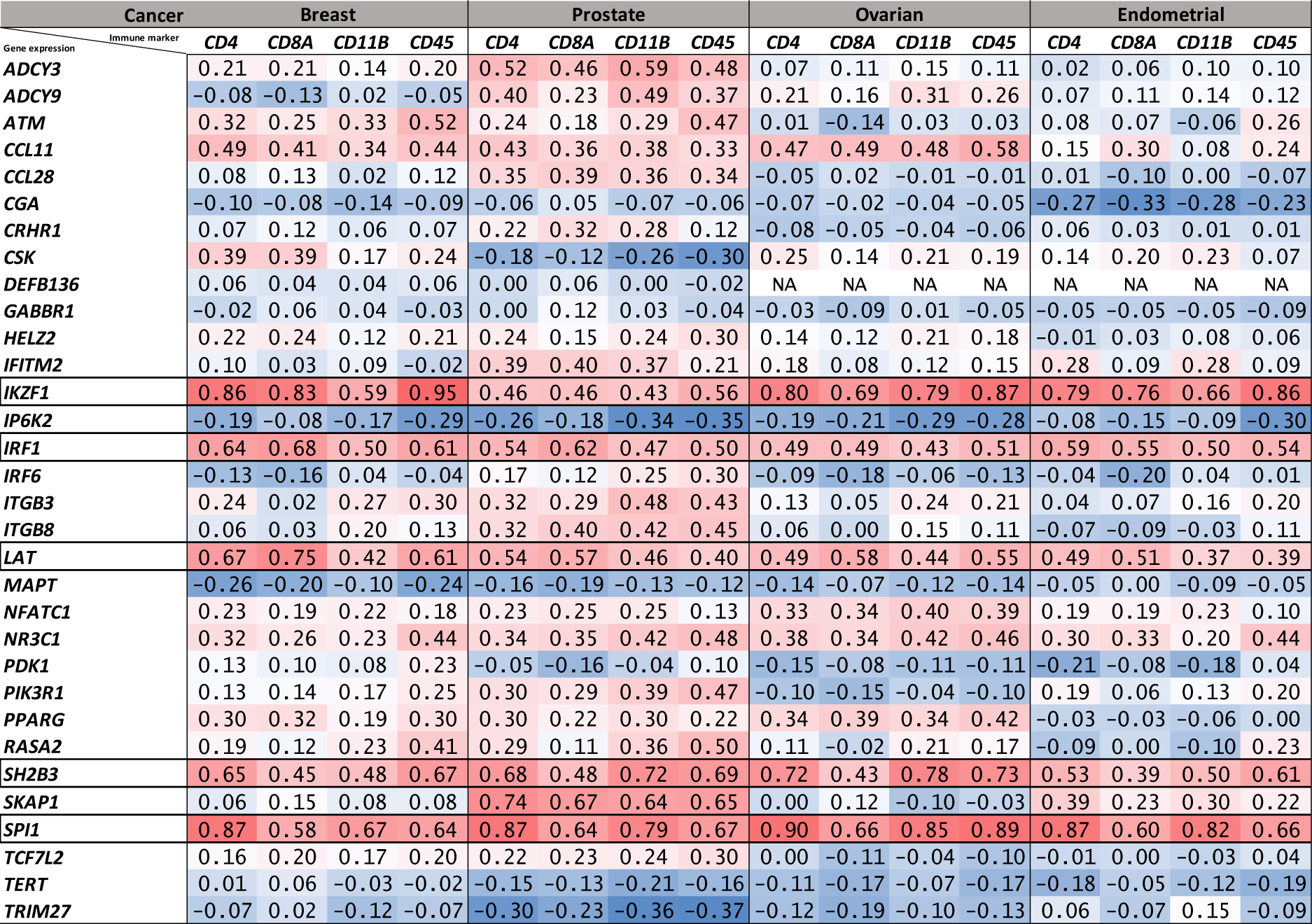
Matrix of expression correlations between immune genes identified by the cross-disorder GWAS meta-analyses and anti-tumor immune infiltrate markers in tumor bulk RNA-Seq data. Numbers shown are Spearman’s rank correlation coefficients (ρ) for correlation in tumor bulk RNA-Seq-based expression levels between the 32 immune-related genes and four T lymphocyte or myeloid cell markers in TCGA breast, prostate, ovarian and endometrial cancers. Each of the 32 genes was the nearest gene for a genome-wide significant lead SNP identified in the pairwise meta-analyses with opposite allelic effects on autoimmune/autoinflammatory disease and cancer. Five genes, with corresponding rows outlined by black borders, were strongly correlated (Spearman’s ρ>0.5) with at least one of the four anti-tumor immune infiltrate markers in all four TCGA cancer cohorts evaluated.

**Figure 2.**
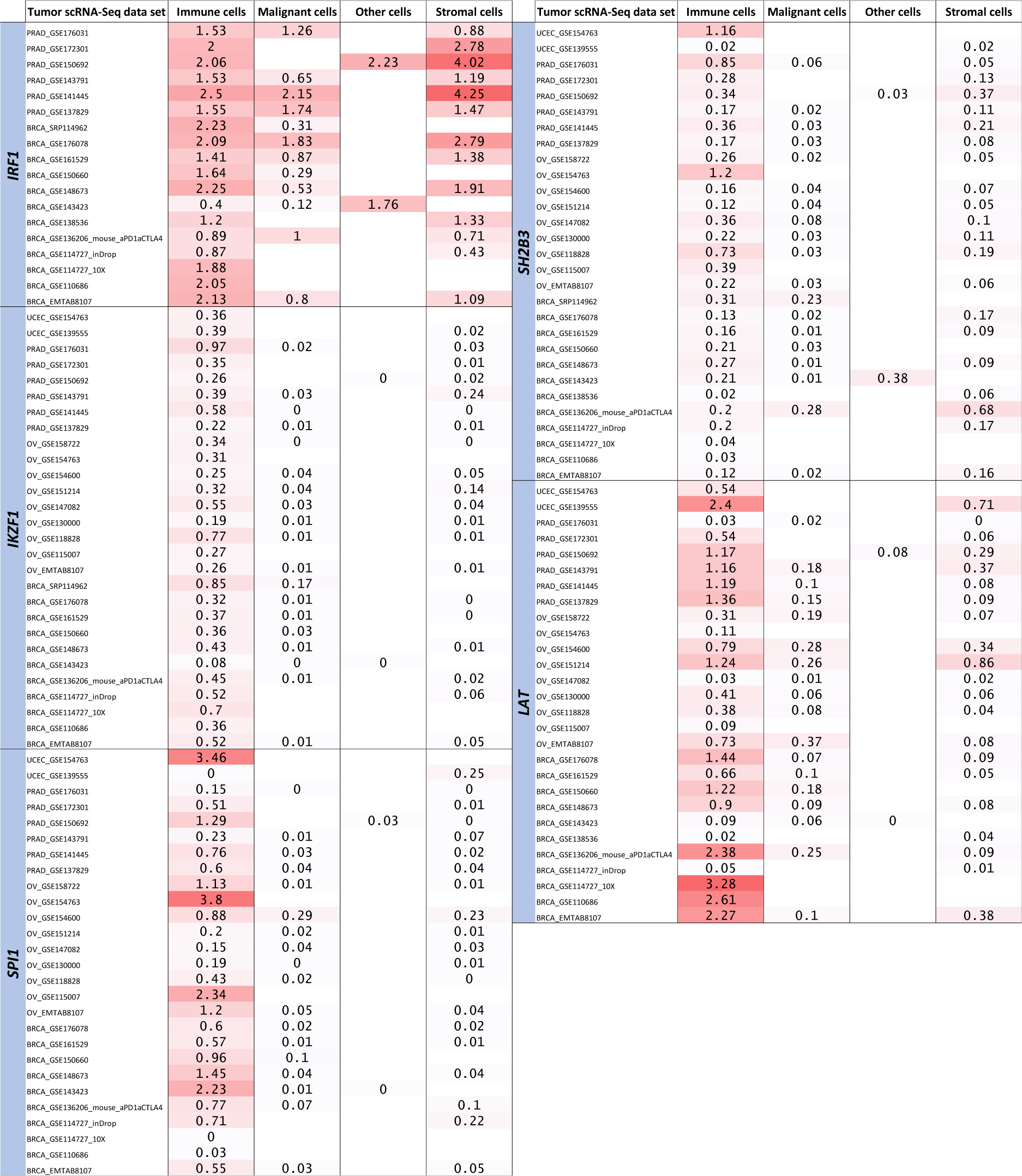
Lineage-wise expression of the five prioritized genes in breast, prostate, ovarian and endometrial tumor single-cell RNA-Seq data. The five genes represent the final targets prioritized by the analytic pipeline from the cross-disorder pairwise meta-analyses, to lead SNP and nearest gene mapping, to the ImmPort, Enrichr and TCGA analyses. The expression of each gene was evaluated across immune, malignant, stromal and other cellular lineages in breast, prostate, ovarian and endometrial tumor single-cell RNA-Seq data sets available in the TISCH2 database. Values shown are in log(transcripts per million/10+1).

**Table 1.** Immune-related genes among the nearest genes for lead SNPs identified in our cross-disorder meta-analyses with opposite allelic effects on autoimmune/autoinflammatory disease and cancer. Full results of the ImmPort and Enrichr analyses are provided in Supplementary Tables 10 and 11.

| Genes | ImmPort | Enrichr | ImmPort category/Enrichr pathway |
| --- | --- | --- | --- |
| *ADCY3* |  | ✓ | Human T-cell leukemia virus 1 infection |
| *ADCY9* |  | ✓ | Human T-cell leukemia virus 1 infection |
| *ATM* |  | ✓ | Human T-cell leukemia virus 1 infection |
| *CCL11* | ✓ |  | Cytokines/Antimicrobials/Chemokines |
| *CCL28* | ✓ |  | Cytokines/Antimicrobials/Chemokines |
| *CGA* | ✓ |  | Cytokines |
| *CRHR1* | ✓ |  | Cytokine receptors |
| *CSK* | ✓ |  | Antimicrobials |
| *DEFB136* | ✓ |  | Antimicrobials |
| *GABBR1* |  | ✓ | Inflammatory Response |
| *HELZ2* |  | ✓ | Interferon Alpha Response |
| *IFITM2* |  | ✓ | Interferon alpha/beta signaling R-HSA-909733/Interferon Alpha Response |
| *IKZF1* |  | ✓ | Development of pulmonary dendritic cells and macrophage subsets WP3892 |
| *IP6K2* |  | ✓ | Interferon alpha/beta signaling R-HSA-909733 |
| *IRF1* | ✓ | ✓ | Antimicrobials/Interferon alpha/beta signaling R-HSA-909733/Interferon Alpha Response |
| *IRF6* |  | ✓ | Interferon alpha/beta signaling R-HSA-909733/Inflammatory Response |
| *ITGB3* |  | ✓ | Inflammatory Response |
| *ITGB8* |  | ✓ | Inflammatory Response |
| *LAT* | ✓ | ✓ | Natural killer cell cytotoxicity/TCR signaling pathway/Modulators of TCR signaling and T cell activation WP5072 |
| *MAPT* | ✓ |  | Antimicrobials |
| *NFATC1* | ✓ | ✓ | Natural killer cell cytotoxicity/TCR signaling pathway/BCR signaling pathway/Human T-cell leukemia virus 1 infection |
| *NR3C1* | ✓ |  | Cytokine receptors |
| *PDK1* | ✓ |  | TCR signaling pathway |
| *PIK3R1* | ✓ | ✓ | Natural killer cell cytotoxicity/TCR signaling pathway/BCR signaling pathway/Modulators of TCR signaling and T cell activation WP5072/Human T-cell leukemia virus 1 infection |
| *PPARG* | ✓ |  | Cytokine receptors/Antimicrobials |
| *RASA2* |  | ✓ | Natural killer cell cytotoxicity/TCR signaling pathway/BCR signaling pathway/Modulators of TCR signaling and T cell activation WP5072/Human T-cell leukemia virus 1 infection |
| *SH2B3* |  | ✓ | TCR signaling pathway/Modulators of TCR signaling and T cell activation WP5072 |
| *SKAP1* |  | ✓ | TCR signaling pathway |
| *SPI1* |  | ✓ | Development of pulmonary dendritic cells and macrophage subsets WP3892/Human T-cell leukemia virus 1 infection |
| *TCF7L2* | ✓ |  | Antimicrobials |
| *TERT* |  | ✓ | Human T-cell leukemia virus 1 infection |
| *TRIM27* | ✓ |  | Antimicrobials |

OpenTargets annotation of the cross-disorder meta-analysis lead SNPs corresponding to the five genes (*IRF1*, *IKZF1*, *SPI1*, *SH2B3* and *LAT*) showed that all five lead SNPs were eQTLs and/or sQTLs for their nearest genes (Table 2). Furthermore, promoter capture Hi-C interactions connected the lead SNPs rs10230978 to *IKZF1* and rs3184504 to *SH2B3*, while the lead SNP rs3740688 was a missense variant (p.Trp262Arg) in *SPI1* (Table 2). We used forest plots to visualize the genetic associations between the cross-disorder meta-analysis lead SNPs corresponding to the five genes and the cancer types and autoimmune/autoinflammatory diseases with which they were associated (Figure 3A). For rs2070721, rs3740688 and rs4788115, the allele that increased cancer risk also increased expression of IRF1, SPI1 and LAT, respectively, in the eQTLGen data (Figure 3B and Supplementary Table 12). For rs10230978 and rs3184504, the allele that increased cancer risk decreased the expression of IKZF1 and SH2B3, respectively (Figure 3B and Supplementary Table 12). Finally, DrugnomeAI predictions indicated that it was highly probable (percentile scores>=94) that the proteins encoded by *IRF1*, *SPI1*, *SH2B3* and *LAT* were targetable via antibody drugs while *IKZF1* received a high probability (percentile score=93) for its protein product likely being targetable by PROTACs (Table 2).

**Figure 3.**
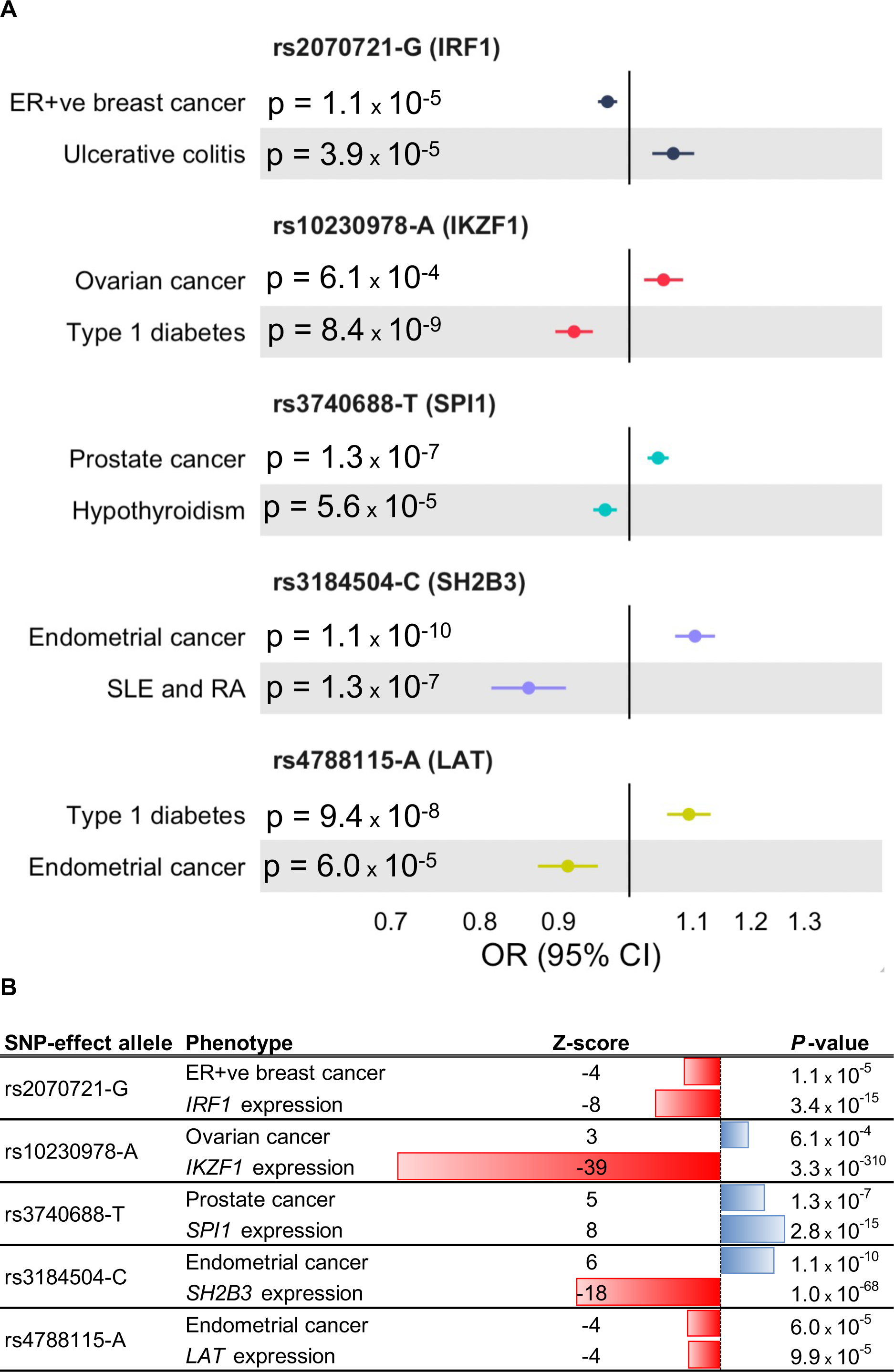
Genetic association forest plots and gene expression associations for the lead SNPs near *IRF1*, *IKZF1*, *SPI1*, *SH2B3* and *LAT*. A. For each lead SNP, the effect allele, nearest gene, P-value, odds ratio (OR) and 95% confidence interval (CI) are shown for the autoimmune/autoinflammatory disease GWAS and cancer GWAS associations. The five genes represent the final targets prioritized by the analytic pipeline from the cross-disorder pairwise meta-analyses, to lead SNP and nearest gene mapping, to the ImmPort, Enrichr and TCGA analyses. For rs3184504-C, the association with SLE is shown while the association with RA is OR=0.93; 95% CI: 0.89—0.95; P=3×10^-7^. **B.** For each lead SNP, the effect allele, Z-score and P-value are shown for the cancer association and for the expression (eQTL) association with the nearest gene.

**Table 2.** OpenTargets evidence linking lead SNPs to nearest genes and DrugnomeAI percentile scores for probability that the protein encoded by a gene is “druggable” for the five genes prioritized by our analytic pipeline (cross-disorder GWAS meta-analyses + nearest gene/immune-related function annotation + tumor bulk and single-cell RNA-seq analyses).

| Lead SNP | Gene | OpenTargets lead SNP-to-gene evidence |  |  |  | DrugnomeAI percentile scores for druggability |  |  |  |  |
| --- | --- | --- | --- | --- | --- | --- | --- | --- | --- | --- |
|  |  | sQTL | eQTL | PCHi-C | VEP | Oncology specific-Antibody | Oncology specific-Small Molecule | Small Molecule | Antibody | PROTAC |
| rs2070721 | *IRF1* | Yes | Yes |  | intronic variant | 92 | 76 | 75 | **95** | 91 |
| rs10230978 | *IKZF1* |  | Yes | Yes |  | 89 | 87 | 85 | 88 | **93** |
| rs3740688 | *SPI1* |  | Yes |  | missense variant | **96** | 82 | 81 | 93 | 88 |
| rs3184504 | *SH2B3* |  | Yes | Yes |  | **94** | 89 | 89 | **96** | **95** |
| rs4788115 | *LAT* | Yes |  |  | intronic variant | **97** | 85 | 84 | **96** | **95** |

Abbreviations: s/eQTL (splicing/expression quantitative trait locus); PCHi-C (promoter capture Hi-C); VEP (variant effect prediction); PROTAC (proteolysis targeting chimera).
Percentile scores >95 (and >90 for PROTAC) are considered “high probability” as per the DrugnomeAI classification.

### Discussion

In this study we have integrated GWAS data sets across seven autoimmune/autoinflammatory diseases and four cancer types and three cancer subtypes spanning over 1.5 million individuals to identify common inherited polymorphisms associated with, and with opposite allelic effects on, cancer and autoimmune/autoinflammatory disease. By bringing together a comprehensive analytic pipeline that included lead SNP-to-nearest gene mapping to link the SNPs to their most probable effector genes, immune-related gene annotation for the nearest genes, correlation of immune-annotated nearest genes with anti-tumor immune response signatures in tumor bulk and single-cell RNA-Seq data, we prioritized five genes: *IRF1*, *IKZF1*, *SPI1*, *SH2B3* and *LAT*. The lead SNP corresponding to each of these genes was an eQTL for the gene and the proteins encoded by the fives gene are potentially druggable candidates. Given all these findings, we highlight the five genes as candidate targets supported by a strong scientific rationale for future in-depth exploration in immuno-oncology drug development programs. Here we discuss each gene in greater detail.

#### IRF1

*IRF1* encodes interferon regulatory factor 1, a transcription factor that binds to interferon-stimulated response elements and plays a vital role in several aspects of the innate and adaptive immune response. IRF1 has been shown to upregulate *CD274* (that encodes PD-L1) expression in the tumor microenvironment and *IRF1* “knock-out” has been shown to render tumor cells more susceptible to CD8+ T cell-mediated killing (30,31). Recently, a peptide inhibitor of IRF1’s interaction with KAT8, a histone acetyltransferase and binding partner of IRF1, has been shown to inhibit PD-L1 expression and improve the anti-tumor immune response both *in vitro* and *in vivo* (32), suggesting that targeting IRF1 or its interactions may represent a promising strategy for cancer immunotherapy. In keeping with these findings, our lead SNP rs2070721-G was associated with reductions in *IRF1* expression and ER-positive breast cancer risk and increased autoimmune/autoinflammatory disease (ulcerative colitis) risk.

#### IKZF1

*IKZF1* encodes an Ikaros family zinc finger transcription factor associated with chromatin remodeling. *IKZF1* expression is specific to the hematopoietic system and serves as a major regulator of lymphocyte differentiation, especially for CD4+ T cells and B cells. IKZF1 functions as a tumor suppressor in acute lymphoblastic leukemia (33). In solid tumors, including prostate cancer, lack of *IKZF1* expression has been implicated as a mechanism for tumor immune evasion (34). Moreover, overexpression of *IKZF1* has been associated with increased recruitment of anti-tumor immune cell infiltrates and sensitivity to anti-PD-1 and anti-CTLA4 therapy (34). The directionality of our genetic epidemiological results were consistent with this biological background: the G allele of our meta-analysis lead SNP rs10230978 which was associated with elevated *IKZF1* expression was protective for ovarian cancer while increasing autoimmunity (type 1 diabetes) risk.

#### SPI1

The *SPI1* proto-oncogene encodes a transcription factor that activates transcription during B-lymphoid and myeloid hematopoiesis. *SPI1* expression is strongly correlated with immune cell infiltration in clear cell renal cell cancer (ccRCC) and non-responders to anti-PD-1 therapy in the CheckMate 09/10/25 ccRCC trials were more likely to overexpress *SPI1* than responders in this cancer type (35). Similarly, in the Imvigor210 anti-PD-1/PD-L1 trial for urothelial cancer, higher expression of *SPI1* and its regulatory targets was associated with significantly lower survival on immunotherapy (36). Biologically and directionally consistent with these findings, in our analyses, the meta-analysis lead SNP rs3740688-T was associated with decreased autoimmune/autoinflammatory disease (Hashimoto’s thyroiditis) risk, increased *SPI1* expression and increased prostate cancer risk.

#### SH2B3

*SH2B3* encodes the lymphocyte adapter protein, LNK. SH2B3 is a negative regulator of several key tyrosine kinases and cytokines, particularly JAK-STAT signaling, and has known tumor suppressor function in acute lymphoblastic leukemia and lung cancer (37,38). SH2B3 is involved in the activation and expansion of CD8+ T cells (39,40). In keeping with these prior biological observations, our analyses indicated that lead SNP rs3184504-C was associated with lower *SH2B3* expression, reduced autoimmune disease (RA and SLE) and higher endometrial cancer risk. In the context of *SH2B3*, it is worth noting that the lead SNP rs318450 is well known to be highly pleiotropic and the allele that decreases cancer risk is associated with increased cardiovascular disease risk probably mediated via an increase in blood pressure (41). Currently approved immune checkpoint inhibitor treatments for cancer are also associated with hypertension and cardiotoxicities (42). Thus, any drug development effort to target SH2B3 may have to find the right balance between inducing cancer suppression and cardiovascular/autoimmune toxicity.

#### LAT

*LAT* encodes the linker protein for activation of T cells. LAT is phosphorylated in response to activation of the T-cell antigen receptor (TCR) and in turn recruits other proteins into major functional complexes at the site of TCR engagement. Controlled modulation of LAT activity either through altered phosphorylation of LAT (43), or by amino acid substitution in the sequence of LAT (44), has been considered as a potential strategy to enhance the efficacy of adoptive immunotherapy in cancer using chimeric antigen receptor or CAR T-cells (45). At the *LAT* locus, we found that lead SNP rs4788115-A was associated with reductions in both *LAT* expression and endometrial cancer risk while increasing autoimmune disease (type 1 diabetes) risk.

Of the 312 unique lead SNP loci with opposite allelic effects on at least one autoimmune/autoinflammatory disease and one cancer type/subtype, we chose to focus on 32 loci where the gene nearest to the lead SNP was clearly immune system related. We then further prioritized 5 of these 32 genes since their tumor expression was strongly correlated with *CD4*, *CD8A*, *CD11B* or *CD45* tumor expression in all four cancers (breast, prostate, ovarian and endometrial). In this context, there are two points worth noting. First, while the corresponding lead SNPs were associated with risk of only one of the four cancers studied (for example, rs10240978-A had opposite effects on ovarian cancer and type 1 diabetes), the five corresponding target genes were correlated with tumor immune cell infiltration marker gene expression in all four cancers. This pattern was consistent with the previously published analyses that uncovered cross-disorder opposite effects at lead SNPs near *CD200*, *CD200R1* and *DOK2* where the germline genetic associations were confined to basal cell carcinoma but the tumor immune gene expression correlations encompassed several additional cancer types (6). Pre-clinical data also showed that anti-CD200R1 antibody treatment had the potential to reverse immunosuppression in multiple cancer types (6). Second, while we prioritized five of the 32 genes the remainder among the 32 also contain genes of potential immuno-oncology interest. For example, lead SNP rs4795899-A (Supplementary Table 5) with nearest gene *CCL11* (Table 1) was associated with protection from Crohn’s disease (P=7.7×10^-7^) and increased risk of ER-negative breast cancer (P=5.9×10^-5^) and was correlated with *CD4*, *CD8A*, *CD11B* and *CD45* expression in TCGA breast tumors with Spearman’s ρ ranging from 0.34 to 0.49 (Figure 1). *CCL11* encodes the major chemokine responsible for eosinophil recruitment and infiltration in the tumor microenvironment (46). DPP4 post-translationally cleaves CCL11 to reduce eosinophil infiltration. DPP4 inhibition (using the anti-diabetic medication, sitagliptin) in a pre-clinical syngenic mouse model of breast cancer has been shown to increase CCL11 levels, eosinophil infiltration and improve tumor control and was synergistic with immune checkpoint inhibition (47).

Gene/protein targets supported by inherited cancer predisposition evidence have been shown to be over twice as likely to be successful at the pre-clinical and clinical phases of oncology drug development (48,49). In the current study, we have provided large-scale germline genetic and tightly coupled immune, somatic and functional genomic evidence to support a deeper evaluation of the proteins encoded by *IRF1*, *IKZF1*, *SPI1*, *SH2B3* and *LAT* as possible targets for cancer immunotherapy.

## Acknowledgments

SPK and his research group receive support from UK Research and Innovation (MR/T043202/2) and the US National Institutes of Health (R01CA259058). JMS and her research group receive support from the US National Institutes of Health (R01CA211574). MPE and his research group receive support from the US National Institutes of Health (RF1AG071170 and R01CA237318). We thank the international consortia that have made the cancer and autoimmune/autoinflammatory disease genome-wide association meta-analysis data sets used in this study publicly available. The results published here are in part based upon data generated by the TCGA Research Network: https://www.cancer.gov/tcga.

## Contributions

SPK conceived and designed the study. JC carried out the main analyses and wrote the first draft of the manuscript. JMS and ME had important roles in study design, data analysis and interpretation of results. JMS, ME and SPK supervised JC and were responsible for data acquisition, funding acquisition and project administration. SPK edited the initial draft of the manuscript. All authors were involved in the review and revision of the final version of the manuscript and approved the final version.

## Data availability

All genome-wide association meta-analysis summary statistics used in this study are publicly available in the GWAS Catalog (https://www.ebi.ac.uk/gwas/home) via accession numbers GCST004132 (Crohn’s disease), GCST004133 (ulcerative colitis), GCST002318 (rheumatoid arthritis), GCST003155 (systemic lupus erythematosus), GCST010571 (autoimmune thyroid disease), GCST005531 (multiple sclerosis), GCST90013445 (type 1 diabetes), GCST004988 (breast cancer), GCST006085 (prostate cancer), GCST004462 (ovarian cancer) and GCST006464 (endometrial cancer).

